# Extending the range of COVID-19 risk factors in a Bayesian network model for personalised risk assessment

**DOI:** 10.1101/2020.10.20.20215814

**Authors:** Georgina Prodhan, Norman Fenton

**Affiliations:** School of Electronic Engineering and Computer Science, Queen Mary, University of London, London, UK

**Keywords:** coronavirus, COVID-19, contact tracing, app, risk, privacy, second wave

## Abstract

A need is emerging for individuals to gauge their own risks of coronavirus infection as it becomes apparent that contact tracing to contain the spread of the virus is not working in many societies. This paper presents an extension of an existing Bayesian network model for an application in which people can add their own personal risk factors to calculate their probability of exposure to the virus and likely severity if they do catch the illness. The data need not be shared with any central authority. In this way, people can become more aware of their individual risks and adjust their behaviour accordingly, as many countries prepare for a second wave of infections or a prolonged pandemic. This has the advantage not only of preserving privacy but also of containing the virus more effectively by allowing users to act without the time lag of waiting to be informed that a contact has been tested and confirmed COVID-19 positive. Through a nuanced assessment of individual risk, it could also release many people from isolation who are judged highly vulnerable using cruder measures, helping to boost economic activity and decrease social isolation without unduly increasing transmission risk. Although much has been written and reported about single risk factors, little has been done to bring these factors together in a user-friendly way to give an overall risk rating. The causal probabilistic model presented here shows the power of Bayesian networks to represent the interplay of multiple, dependent variables and to predict outcomes. The network, designed for use in the UK, is built using detailed data from government and health authorities and the latest research, and is capable of dynamic updates as new information becomes available. The focus of the paper is on the extended set of risk factors.

## I. Introduction

Like many countries, Britain is bracing for a second wave of coronavirus. Local lockdowns are being reimposed in areas of England, which suffered the highest levels of excess mortality in Europe (Office for National Statistics, 2020), even as business are encouraged to stay open, individuals urged to go back to work in offices, and schools go back for the new academic year. Mitigating measures such as the compulsory wearing of face masks in confined public spaces have recently been imposed. But a digital contact-tracing system that was supposed to be the basis for a lifting of restrictions on movement and social activity has yet to prove itself effective. The UK government has just launched a revamped version of its coronavirus app. It has also been reported to be considering a feature that allows people to use personal information to calculate their own risk score (Smyth & Wright, 2020). The model presented here could be the basis for just such an app. The location of new outbreaks could be identified simply by collecting data on users’ location (via GPS, not the more invasive Bluetooth) and likelihood of infection.

This approach to containing the virus relies largely on individuals taking responsibility for their own health and that of those around them. This may be thought to be a weakness. But in fact, in all but the most authoritarian regimes, a sense of social responsibility is essential if rules on social distancing, face masks, hand-washing and other measures are to be effective. Globally, experience of past pan- and epidemics including the 2009 swine flu, SARS, the 2014-2016 Ebola outbreak and various outbreaks of bird flu have shown that people’s behaviour – based on risk perception - can have a profound influence on the spread of infectious diseases (Funk, et al., 2009).

There is evidence that the model presented here could be effective in Britain (although it is, of course, adaptable to any country in the world). A study in 10 countries that was published in May (Dryhurst, et al., 2020) found that the UK had the highest public risk perception of COVID-19 in the group. The COVID Symptom Study, an epidemiological research app that invites users to report any symptoms and share details of their health lifestyle, has more than 4 million contributors, mostly in the UK (Zoe, 2020). And, according to a poll of 2,254 UK residents carried out in in May by King’s College and Ipsos MORI (Allington, et al., 2020), the majority have been going beyond compliance with government advice, for example by staying at home for long periods. The poll also found that most people would obey a contact-tracing app’s recommendation to self-isolate – showing the potential utility of an app that relies on voluntary participation. However, the majority did not trust the government to keep their data safe, and almost half were sceptical about the ability of such an app to limit the spread of coronavirus.

This paper is part of a set of studies conducted by a multidisciplinary team of researchers who argue that contact tracing alone is unlikely to contain a high-prevalence, contagious disease such as COVID-19. It builds on the work most recently of (Fenton, et al., 2020), which describes details of a Bayesian model to compute the probability that an individual has COVID-19 – whether symptomatic or asymptomatic – or is likely to catch it. The model presented here expands substantially on that web of relevant risk factors, bringing in ethnicity, religion, occupation and housing conditions, and refining other factors such as age, obesity and underlying medical conditions. It also incorporates concurrent work on an extended set of symptoms described in (Butcher, 2020).

Results of running the model are consistent with what is known about the prevalence and severity of COVID-19, and the vulnerability of different groups. This demonstrates its ability to predict risks for individuals, even if relatively little is known about them. For example:

- Entering the observation that someone is Black or non-Chinese Asian results in a raised probability of having severe COVID-19 in the 55-64 age group and an equal probability in every other age group. This is consistent with observed data (Office for National Statistics, 2020). The opposite is true, however, when comparing ethnic groups without conditioning on age - an interesting paradox that will be discussed below.
- Entering the observation that someone is aged under 16 reduces the probability of having severe COVID-19 to virtually zero but raises the probability of having the disease asymptomatically. This is consistent with observed data and an important factor to watch as schools reopen (Centers for Disease Control and Prevention, 2020). Adding the observation “student/pupil” raises the probability of eventual COVID-19.
- The probability of infection takes precedence over background risks – living in an overcrowded household is more dangerous than being obese. This is logical, since nobody can have COVID-19 without becoming infected, no matter what their vulnerabilities. It shows the power of mitigating measures, and cautions against overemphasising personal risks such as obesity or smoking.

## II. CONTEXT AND RELATED WORK

Countries around the world have introduced contact-tracing apps in attempts to contain a second wave of coronavirus as lockdowns are eased. Contact tracing using mobile technology has become a particular focus during the current epidemic because COVID-19 presents the challenge that the majority of transmission is believed to occur before symptoms of the disease show. This differentiates it from ebola, smallpox or the 2002 SARS-CoV-1, and makes it vital to reduce the time from symptoms appearing in a person to isolation of that person’s contacts. Recent studies including (Ferretti, et al., 2020) and (Kretzschmar, et al., 2020) suggest that the time from confirmation of a COVID-19 case to quarantining of contacts needs to be under three days to bring the rate of reproduction below 1, thus containing the virus’s spread. Such a time frame is generally beyond the capabilities of manual tracers, especially for an epidemic of the current scale. The (Ferretti, et al., 2020) study, which examined early epidemic data from China, Singapore and the Diamond Princess cruise ship, concluded that near-instantaneous contact tracing via a mobile phone app, in conjunction with other measures, could contain the virus.

While adoption of mobile contact-tracing apps has been relatively high and has been credited with helping to flatten the curve in some East Asian countries, the same conditions do not apply in much of the West (Huang, et al., 2020). In China, using a traffic-light health-code system embedded in the extremely popular WeChat and Alipay platforms is practically mandatory for freedom of movement. In South Korea, which swiftly contained the first outbreak (although it is now battling a second), authority had already been given to disease-prevention authorities to override some privacy laws during the 2015 MERS outbreak (Park, et al., 2020). Crucially, the country also quickly developed large-scale testing infrastructure. In Singapore, the first country to deploy a national coronavirus-tracing app, a wearable token is being rolled out to address weaknesses in the existing mobile-phone technology – and has sparked a rare backlash against the government, with accusations it is on the way to becoming a surveillance state (Asher, 2020).

Among Western countries, adoption of government-endorsed mobile contract-tracing apps has been relatively high in Australia, Germany and Italy, with rates of around 22%, 14% and 7% respectively by mid-July (Chan, 2020). But none of these countries can show or is willing to claim the effectiveness of the app in limiting the spread of the virus. And even the countries with the highest adoption are still well below the threshold of 56% of the population, or 80% of smartphone users, that experts consider the minimum for the technology to have a chance of being effective (Hinch, et al., 2020). Privacy concerns are largely to blame, along with technical glitches and discomfort with digital technology among some groups, especially older people.

Some privacy-preserving contact-tracing apps are already on the market or in development. A few stand out:

- Private Kit: Safe Paths, developed at Massachusetts Institute of Technology. This is an open-source technology that that provides individual users information on their interaction with COVID-19, and allows them to share their location trails with health officials if they test positive for the disease (Rasker, et al., 2020). The app is available for iOS and Android.
- COVID-19 Watch from Stanford University. This is an early-warning system using Bluetooth, which allows an infected person to send an anonymous alert to others they may have infected, if they are also users of the app (Abate, 2020). It is currently in testing.
- CoEpi. This is a community-based epidemiological tool that uses Bluetooth technology and is based on voluntary symptom-sharing, including before confirmed test results (CoEpi, 2020). It is currently in development.

All of these apps and proposed apps address privacy concerns, and CoEpi also potentially collapses the time from infection to isolation of a contact by promoting symptom-sharing before test results. They all deal exclusively with infection risks, however, and none incorporates background risk to give a personalised risk assessment in the way that the model presented here does.

I searched the preprint servers medRxiv, bioRxiv and arXiv as well as Google Scholar and LitCovid for Bayesian approaches to risk assessment and contact tracing. I found several interesting papers on transmission dynamics and mitigation measures such as physical distancing, face masks and eye coverings. An early study of Chinese data formulated an equation to determine personal risk based on age, sex and comorbidities (Caramelo, et al., 2020). It used a naïve Bayesian approach, assuming that all the risk variables were independent of one another. An interesting French study modelled individual risk as a potential decision-making aid to relaxing lockdown restrictions on low-risk populations (Evgeniou, et al., 2020). It used standard statistical and machine-learning methods such as logistic regression and random forest. Another, Italian, paper aimed to develop a personal risk score for infection (Orlando, et al., 2020). However, it only measured association, not causality, which was evident in its finding that underlying medical conditions could influence the risk of infection. In causal Bayesian terms, this would be a category mistake as the most vulnerable person in the world could not become infected with coronavirus if they did not come into contact with it. I found nothing that used a Bayesian analysis for personal risk prediction and containment, apart from the work on which this project is based (Fenton, et al., 2020).

## III. METHODOLOGY

The model presented is a Bayesian Network, a causal probabilistic model that predicts outcomes – in this case, current and projected COVID-19 status and severity of disease – using a combination of actual observations and expert calculations. It is built on AgenaRisk Bayesian network software. A simplified schema of the network is shown in Fig. 1, and a more detailed version can be found in the Appendix. The model, named “CombinedModelForContactTracing” can be accessed using this link http://www.eecs.qmul.ac.uk/~norman/Models/ and can be run using trial software from agenarisk.com.

**Fig. 1.**
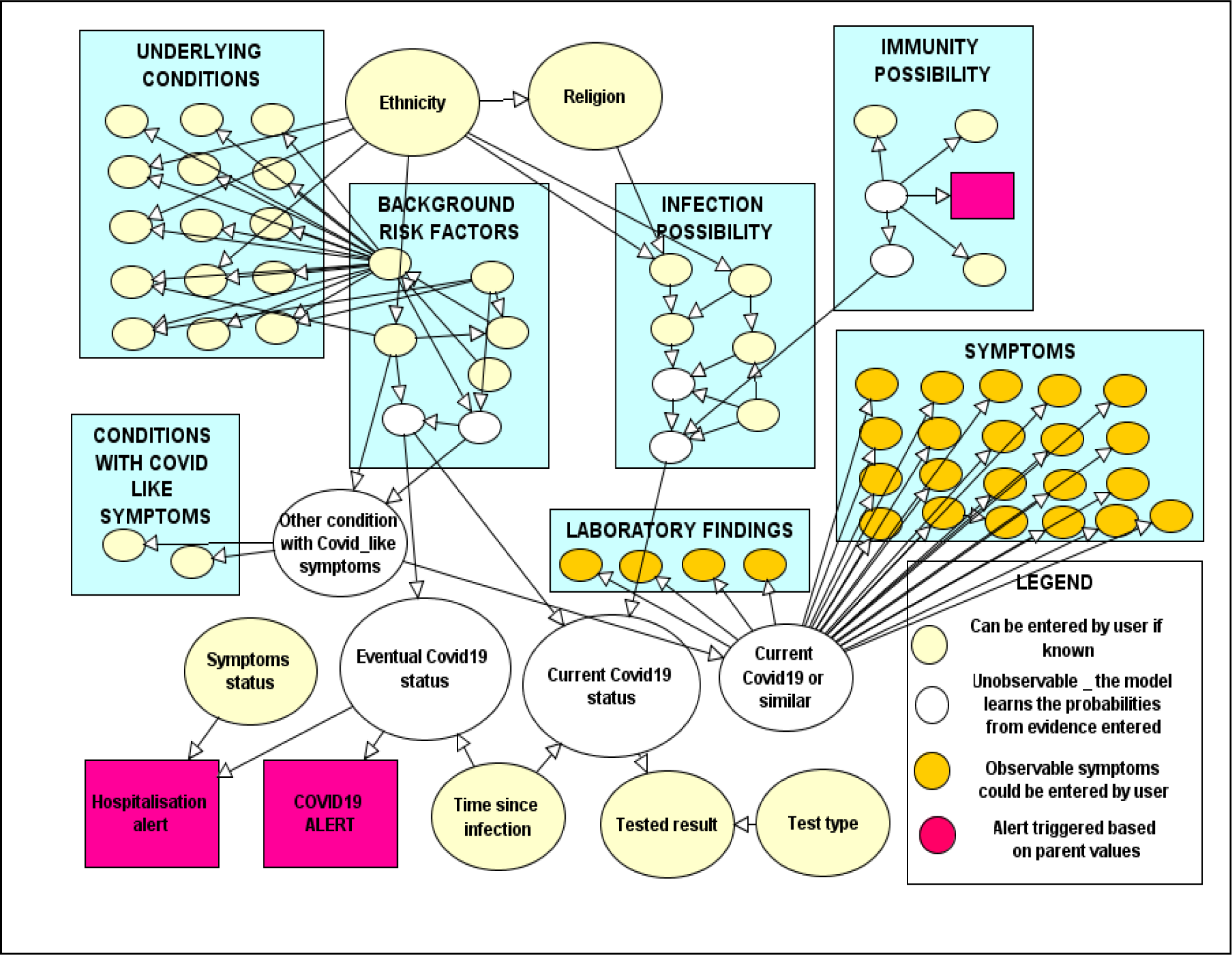
A simplified version of the Bayesian network for COVID-19 risk assessment.

The network is initially populated with prior probabilities for the general population – e.g., a person in the UK has a 49.4% chance of being male, a 23.4% chance of being obese and a 16% chance of being a smoker. Together with other factors including infection opportunities, and immunity possibility, these resulted in a 0.2% chance of eventually having COVID-19, at the time of writing.

Bayesian networks are known to be able to solve problems that traditional statistics cannot, notably cases of confounders and colliders – in which associated variables are causally related only through a common cause or a common effect. A Bayesian approach is well-suited for the current situation, in which it is crucial to understand the relationships of multiple dependent variables, and where observational data have to be gleaned opportunistically as controlled experimentation is impossible.

Prime examples of confounding variables surround the issue of ethnicity. Several large studies including (The OpenSAFELY Collaborative, 2020) and (Office for National Statistics, 2020) found a significantly higher risk of death from COVID-19 among Black or South Asian people than among White people. But by looking only at association or correlation, their methods were by definition unable to discover the mechanisms by which this occurred. Even after it said it had adjusted for age, region, population density, area deprivation, household composition, socio-economic position, education, household tenure, multigenerational households and occupation, the ONS still found that Black males had twice the risk of White males, which it found to be “unexplained”.

In the Bayesian network presented here, it can be clearly seen that the relationship between ethnicity and COVID-19 death is in fact confounded by multiple variables: Black and South Asian people are more likely to be frontline healthcare workers, live in overcrowded households or have sickle cell disease, for example, than White people. The healthcare worker case is illustrated in Fig. 2. All of these are factors that increase the risk of catching COVID-19 or may lead to a more severe form of the disease. They confound the relationship between ethnicity and COVID-19 outcomes.

**Fig. 2.**
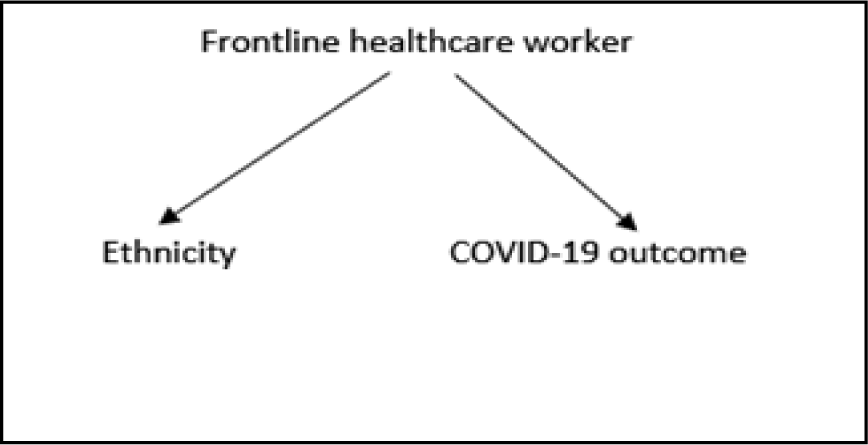
The variable “frontline healthcare worker” confounds the relationship between ethnicity and COVID-19 outcome

Potential collider variables are also abundant in the current COVID-19 situation because of the nature of the studies that have been carried out. Observational data cannot be randomly sampled and controlled experiments cannot be done so almost all the data available come from hospital populations, individuals who have been tested or people voluntarily taking part in studies. These are not representative of the population: hospital patients are already frail, tested individuals are already exposed or symptomatic, and voluntary participants may tend to be health-conscious, well-educated, tech-savvy or hypochondriac. As such, any study conducted on one of these populations is already conditioned on a collider variable such as “frail”, “vulnerable” (e.g. frontline healthcare worker) or “tech-savvy” (i.e. likely to be younger). A UK government community infection pilot study (Office for National Statistics, 2020), which conducts regular tests on a large number of randomly selected participants, is beginning to redress this situation to some extent.

Observing associations between other factors in such pre-selected conditions can result in misleading conclusions. Many of the underlying medical conditions associated with more severe COVID-19 may bear no causal relation to severity of COVID-19 but simply have been observed in hospitalized patients with severe COVID-19 because those conditions make them more likely to be hospitalised in the first place. For example, it is not completely clear how heart disease may influence COVID-19 outcome, but by conditioning studies on the collider variable “hospitalised” we are confining our observations to a population already more likely to have heart disease or a whole range of other co-morbidities, and ignoring people who have those conditions combined with mild COVID-19 but remain out of hospital. In this way, an exaggerated or even a false association can appear between the variables “heart disease” and “severe COVID-19”, as seen in Fig. 3. The U.S. Centers for Disease Control and Prevention has noted that only 6% of the COVID-19 deaths so far have mentioned COVID-19 as the sole cause of death (National Center for Health Statistics, 2020).

**Fig. 3.**
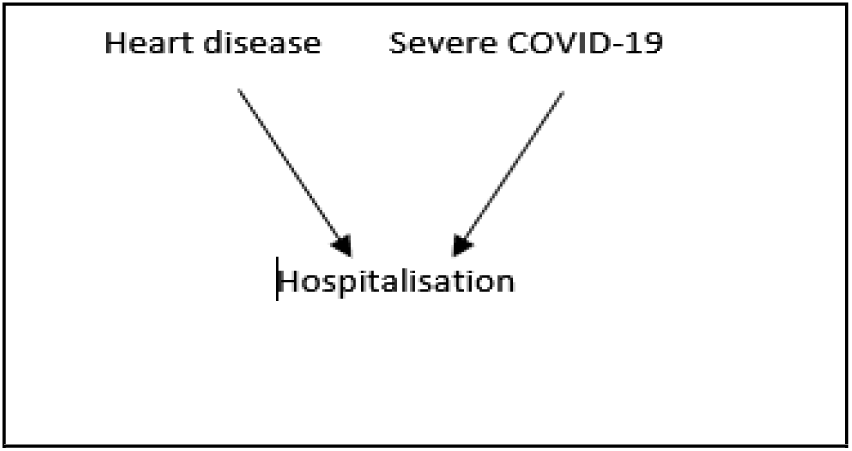
Heart disease and severe COVID-19 collide on the variable “hospitalization”

In the current version of the model, I have included the underlying medical conditions that best expert advice considers linked with higher risk of severe COVID-19 because of a lack of further evidence to causally explain them or explain them away. But the list has changed many times during the making of the model and may change again.

It is worth noting that even though male sex is widely observed and accepted as being associated with higher risk, this has not yet been convincingly explained in a causal way and may yet turn out to be a relationship that is confounded by other factors. It has been suggested that the hormone oestrogen (which men have in smaller amounts) is a protective factor. A new study argues that females may mount a more robust immune response than males, even in older age (Takahashi, et al., 2020). But research is still at an early stage. Again, a Bayesian model can easily adapt to new knowledge, for example by creating a new node “oestrogen level” and drawing edges connecting this to both males and females, with different prior probabilities.

## IV. MODEL ARCHITECTURE

The Bayesian network is an acyclic directed graph that consists of probability nodes connected by edges representing conditional dependencies. Inside each probability node is a probability table. Some of these are simple, e.g. for sex or for age, which are not conditioned on any other factors – the nodes have no “parents” – as seen in Fig. 4. In other cases, they are more complex, e.g. the diabetes node probability table is conditioned on underlying medical condition and ethnicity – as seen in Fig. 5.

**Fig. 4.**
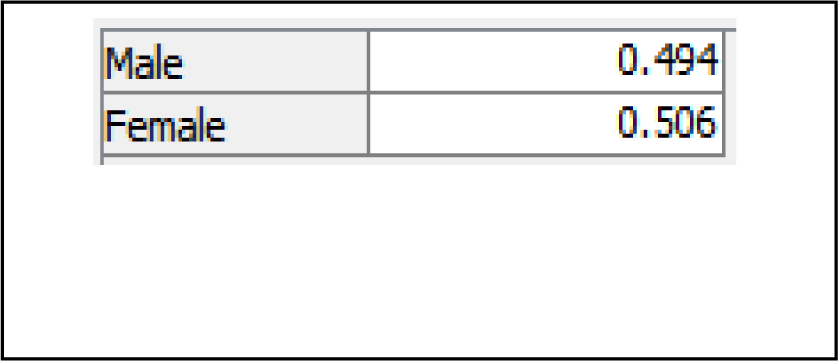
Node probability table for “sex”.

**Fig. 5.**
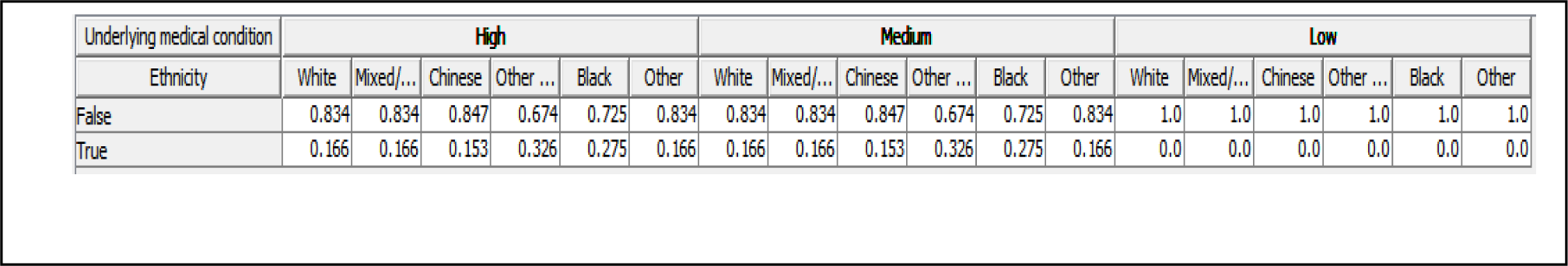
Node probability table for “diabetes”.

The structure of the network, inherited from (Fenton, et al., 2020), consists of four main areas that feed into the calculation of probability of COVID-19: background risk factors, infection possibility, immunity possibility, and symptoms. I developed the areas of background risk factors and infection possibility. My colleague Rachel Butcher developed the symptoms area.

Entering an observation in any of the nodes will change the probabilities of all the variables that are connected to it, whether in the direction of influence or by backward inference. For example, entering an observation that a person is aged 75 or older will raise the probability of currently having severe COVID-19 from 0.01% to 0.02% and of eventual severe COVID-19 from 0.03% to 0.06%. But it will also raise the probability of being White from 84.8% to 95.9% (because White people live longer), it will raise the probability of being Christian from 58.7% to 62.6% (because more White people are Christian) and it will decrease the probability of being morbidly obese from 2.8% to 1.5% (because some of the morbidly obese will have died by that age, and elderly people tend to eat less). As many facts as are known can be entered - the more observations, the more accurate the prediction.

A few of these nodes and how their probability tables were arrived at will be discussed below. Details of all the node probability tables that I developed are in the Appendix, along with their data sources.

### A. Background risk factors

At the time of writing, there was broad consensus that age, sex, obesity and certain underlying medical conditions were important risk factors for developing more severe forms or dying of COVID-19 (Verity, et al., 2020) (Gebhard, et al., 2020) (Petrakis, et al., 2020) (Clark, et al., 2020) (Blackshaw, et al., 2020). The main difficulty arose in how to represent the interplay of the various factors, given that age, sex and obesity were themselves interrelated and in addition could all influence the probability of underlying medical conditions. It would be impossible to give accurate probabilities for each combination of these (e.g. probability of having an underlying medical condition for an overweight woman aged 35-54, and then the probability of an underlying medical condition for that same woman if she were normal/underweight, obese or morbidly obese, and so on). In fact, for seven age categories, two sex categories and four obesity categories there would be 56 combinations of factors that could each lead to a high, medium or low probability of having an underlying medical condition. Clearly, it would be over-engineering the model to give a value to each of these.

Instead, I drew links between nodes where I did have accurate figures. For example, there were reliable data for obesity by age group and sex so I made age and sex parent nodes of obesity. I made particular underlying medical conditions child nodes of the general underlying medical conditions node, with age, obesity and sex as parent nodes of those individual conditions where their influence was known to be strong.

To capture the complexity of the influence of ethnicity on COVID-19 risk, the node “ethnicity” was placed outside the background factors section of the model as a parent of other factors such as age, underlying medical condition and overcrowded housing (an infection factor).

Despite a flurry of excitement around a French study released in June that proposed smoking may have a protective effect against developing COVID-19 symptoms (Miyara, et al., 2020), this was soon dismissed by expert consensus, and the direct effects of smoking on COVID-19, if any, are still unknown (Zyl-Smit, et al., 2020) (Fenton, 2020). Smoking was therefore included in the model as a parent node of “underlying medical condition” and also of chronic obstructive pulmonary disease (COPD).

### B. Infection possibility

Occupation was the single most important factor in determining the probability of exposure to the virus, with frontline healthcare workers clearly the most exposed. South Asian and Black people are overrepresented in the UK’s National Health Service and therefore especially in early COVID-19 deaths, when personal protective equipment (PPE) was scarce.

Living in multi-generational households is also a risk factor, especially in cases where a middle generation goes out to work and lives with elderly relatives and with children who may socialise more outside the home. This is more common in South Asian communities but is also becoming generally more popular as elderly people move in with their children, while adult offspring move back or stay in their parental home for economic reasons. I drew a link from ethnicity to overcrowded household, as data exist for overcrowding by ethnicity. I also drew a link from religion to overcrowded household, mainly to account for the large multi-generational households of Britain’s strictly Orthodox Haredi population, which was not captured by ethnicity (Judah, 2020).

## V. SELECTED PROBABILITY NODES

### A. Ethnicity

The modelling of ethnicity was one of the most complex parts of building the network. A whirlwind of media and statistical reports was published in Britain and the United States regarding the higher vulnerability of non-White ethnic groups to severe COVID-19 symptoms, hospitalisation and death in the wake of the killing of George Floyd and subsequent surge in support for the Black Lives Matter movement that occurred while I was working on the model. No substantial evidence has as yet emerged of genetic factors that would explain a racial bias in the spread of the disease. Genomics England and the University of Edinburgh have launched a nationwide study to understand how a person’s genetic makeup could influence how they react to the virus. (Genomics England, 2020). This is still in progress.

The media and political storm did cause a lot of data to be published analysing COVID-19 statistics by ethnicity, which was very useful in the building of this model. Therefore, I was able to represent the relationship between ethnicity and severity of COVID-19 risks with some accuracy through a greater tendency of ethnic minorities in the UK to live in overcrowded housing, suffer from certain health conditions such as diabetes, and work on the healthcare frontline. I have reproduced below a visualization of the complexity of the situation by (Pareek, et al., 2020) in Fig. 6.

**Fig. 6.**
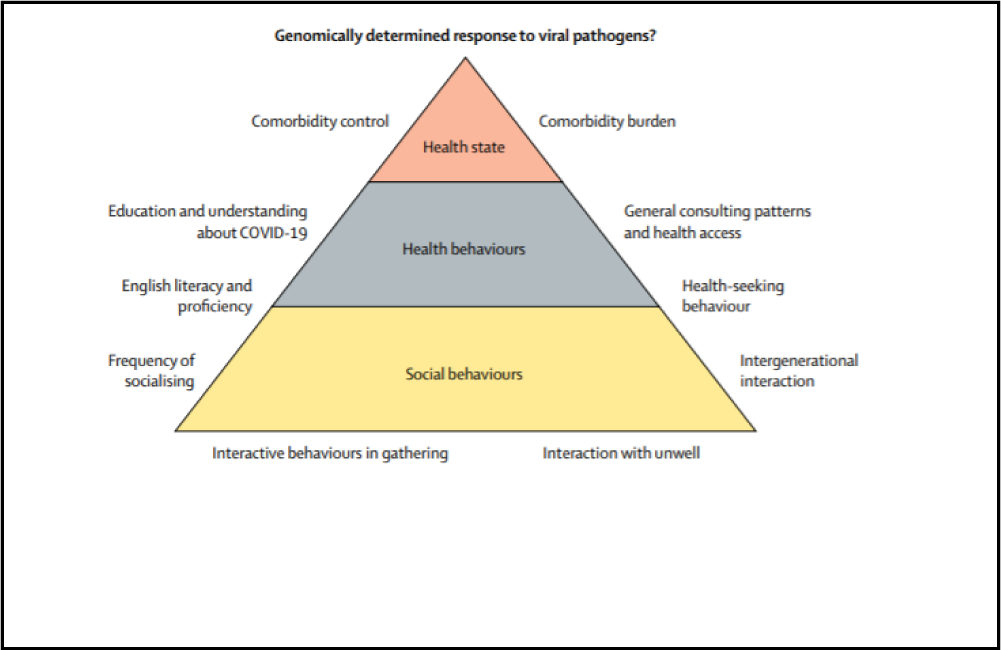
The potential interaction of ethnicity-related factors on SARS-CoV-2 infection likelihood and COVID-19 outcomes (Pareek, et al., 2020)

Even with only a few of these important factors captured, the model is capable of assessing that a White, female nurse of normal weight living in shared accommodation is at higher risk than a Black, obese male office clerk working from home, where he lives with his nuclear family – even though being Black and obese may overall be likely to make for higher risk in the absence of other knowledge. A fragment of the model showing the edges connecting ethnicity to other nodes is shown in Fig. 7. Note that some of these factors – short lifespan, overcrowded household, risky occupation – allow for interventions to make matters more equitable. Ethnicity is of course not a variable that can be changed.

**Fig. 7.**
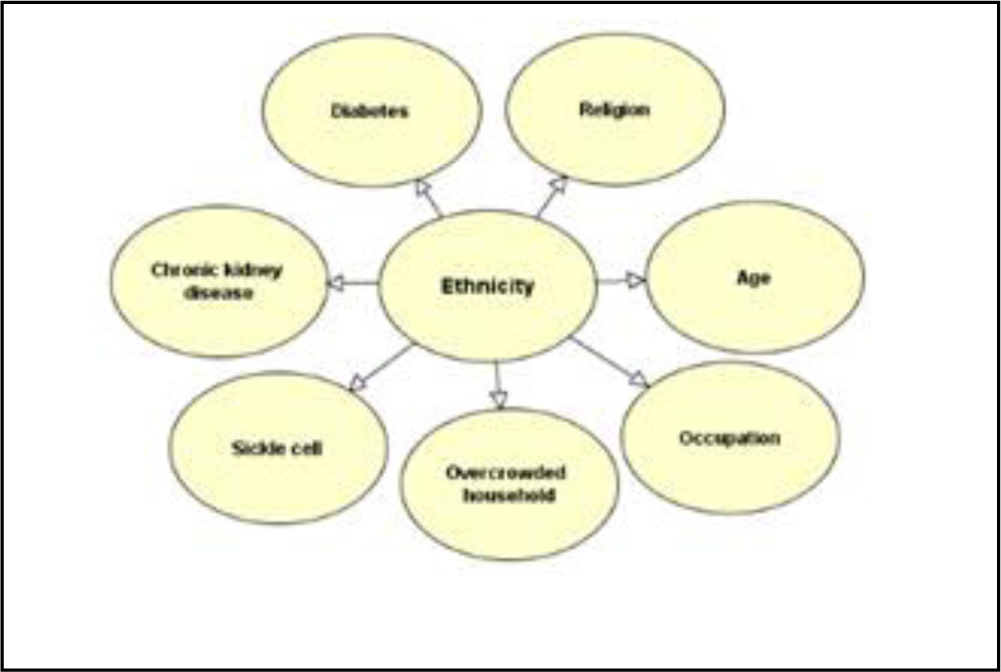
Model fragment “ethnicity”.

The connection between age and ethnicity is an important one. It explains the paradoxical situation that Black and non-Chinese Asian people have a lower risk of developing severe COVID-19 when considered as whole populations – in apparent contradiction of the news headlines – but they do have an equal or increased risk when considered age group by age group. This is an example of Simpson’s Paradox, in which a characteristic of a larger group can disappear or be reversed when considering subgroups one by one. The situation is summarized in Table 1.

**TABLE 1.** PROBABILITY OF EVENTUAL SEVERE COVID-19 ILLNESS BY ETHNICITY AND AGE.

| AGE | White | Asian* | Black |
| --- | --- | --- | --- |
| Unobserved | 0.03% | 0.02% | 0.02% |
| 75+ | 0.06% | 0.06% | 0.06% |
| 65-74 | 0.05% | 0.05% | 0.05% |
| 55-64 | 0.03% | 0.04% | 0.04% |
| 35-54 | 0.02% | 0.02% | 0.02% |
| 25-34 | 0.02% | 0.02% | 0.02% |
| 16-24 | 0.01% | 0.01% | 0.01% |
| <16 | 0.01% | 0.01% | 0.01% |

\*excluding Chinese

The reason for this is simply that White people, who make up the majority of the UK’s population, tend to live longer than other ethnic groups. Age is also the single most important determinant of COVID-19 severity. Taken overall, the UK’s Black and non-Chinese Asian populations are relatively young, with only a small proportion living long enough to fall into higher-risk age categories. Considering the sub-categories of age group rather than the whole population is therefore the right way to analyse the situation, comparing like with like. This is a good validation of the model. The effect is similar but more pronounced in the United States, where infection rates are still significantly higher than in Britain (Mackenzie, 2020).

### B. Obesity

Certain underlying medical conditions are widely believed by experts to contribute to the risk of developing more severe COVID-19 symptoms or dying of the disease. Some large-cohort studies of hospital patients have analysed factors associated with COVID-19 death (The OpenSAFELY Collaborative, 2020) (Emami, et al., 2020) (Docherty, et al., 2020). These are observational studies and, as such, do not establish causal relationships. Nonetheless, they represent best expert opinion at the time of writing. Many experts believe that, broadly, underlying medical conditions weaken the immune system and thus limit the body’s ability to fight off infection or illness.

One such condition is obesity, and in late July the UK government launched a campaign to fight it, arguing that people could make themselves less vulnerable to a second wave of coronavirus by losing weight. Public Health England published a long report on the topic, which synthesised a large amount of useful information but was flawed in many ways. For example, it argued:

“Evidence suggests excess weight is associated with an increased risk of the following for COVID-19: a positive test, hospitalisation, advanced levels of treatment (including mechanical ventilation or admission to intensive or critical care) and death.” (Blackshaw, et al., 2020)

It is unclear why it would be worth noting that excess weight is associated with a positive test since, as the report itself acknowledges, obesity does not increase the chance of infection. Secondly, the report highlighted the fact that 31.3% of patients critically ill in intensive care units (ICUs) were obese, with a body mass index (BMI) of 30 or over, compared with 28.9% of the population, while 7.9% were morbidly obese, with a BMI of 40 or over, compared with 2.9% of the general population. Both were adjusted for age and sex in unspecified ways. Aside from the fact that the difference in the case of simple obesity is very small, this gives a misleading impression of the risks of obesity when reading the results in the causal direction – from obesity to intensive care. In my model, entering the observation “severe” in the “current COVID status” node also raises the probabilities of being morbidly obese or obese, as seen in Table 2. (The data used differ slightly from those in the PHE report.)

**TABLE 2.** PROBABILITY OF OBESITY GIVEN SEVERE COVID-19.

|  | Prior probability | Observe current COVID-19 status severe |
| --- | --- | --- |
| Morbidly obese | 2.846% | 3.465% |
| Obese | 23.353% | 27.836% |
| Overweight | 31.05% | 33.947% |
| Normal/underweight | 42.75% | 34.752% |

However, reading the data in the direction of influence by clearing the observation “severe COVID-19” and observing obesity and morbid obesity in turn has no perceptible effect on COVID-19 status measured to two decimal places, as seen in Table 3.

**TABLE 3.** PROBABILITY OF EVENTUAL COVID-19 GIVEN OBESITY.

|  | Prior probability | Observe obese | Observe morbidly obese |
| --- | --- | --- | --- |
| COVID-19 status severe | 0.03% | 0.03% | 0.03% |
| COVID-19 status mild | 0.11% | 0.11% | 0.11% |
| COVID-19 status asymptomatic | 0.07% | 0.06% | 0.06% |
| COVID-19 status none | 99.8% | 99.8% | 99.8% |

This shows that backward inference is not a guide to effectiveness of intervention and, while losing weight is certainly good health advice for many people, an anti-obesity drive may be a waste of public time and money as a measure to fight the effects of any second coronavirus wave.

I included obesity in the model as a parent of “underlying medical condition” as, like smoking, it is a predictor of several conditions. I did not include it as a parent of “risk factors” in its own right.

### C. Occupation

Occupation is a key determinant of exposure to coronavirus. Frontline healthcare workers are at significantly higher risk because they are exposed to many people, and those people are far more likely than the general population to be ill with COVID-19. Other essential workers who have contact with others at work or who have to travel to their place of work on public transport are also significantly more exposed than the general population. The UK’s Office for National Statistics has analysed occupations with the highest potential exposure (Office for National Statistics, 2020). In some cases, numbers are small and so this is a work in progress. Risky occupations outside healthcare included bus drivers, care home workers and home carers, security guards, primary and nursery teachers, police officers, opticians and pharmacists, plumbers, vets and undertakers. I included students/pupils as an occupational category in their own right, as COVID-19 outbreaks began to appear on campuses and in schools at the start of the new academic term.

It is envisaged that users of the app would have a multiple-choice question about their occupation, which would yield a high, low or medium risk category. The actual list of risky occupations could be adjusted as more research became available.

The ONS analysed those employed in these areas by sex and ethnicity. Some occupations were skewed towards women (carers and teachers), some towards South Asians (pharmacists and opticians) and some towards white males (police officers and plumbers). Overall, there seemed no great imbalance in the chances of belonging to one of these medium-risk occupational groups so I did not consider ethnicity or sex as parents of those.

I did, however, do so for frontline health workers, using data from the NHS, which has critical mass with 1.24 million employees, making it the biggest employer in the UK, and where Black and south Asian employees are overrepresented.

## VI. SCENARIOS

A few fictional cases illustrate the power of this model over simple contact-tracing exposure models, and also its ability to give a risk assessment far more useful to individuals than the general government guidance.

### A. The unemployed White male

Fred is a 55-year-old unemployed White man. He is obese and he smokes but he has no known medical conditions. He lives alone and has minimal interactions with people outside the home. He describes his religion as “none”. He has not knowingly had any contact with people with COVID-19 or COVID-19 symptoms. He worries about his weight and his smoking but in fact, the model predicts, his risk of eventual severe COVID-19 is lower than average and he is strongly predicted not to get COVID-19 at all, with a probability of 99.85%.

### B. The retired female country-dweller

Katie is a 79-year-old, white, retired lawyer. She has a touch of Parkinson’s Disease but is of a healthy weight and has never smoked. She lives in a small village with her husband and has had minimal contact with other people since the lockdown. She is Catholic and goes once a week to church, where she socially distances. Katie is worried about her vulnerability, mainly because of her age, and follows government advice to shield at home, not even venturing out for walks. The model calculates her probability of eventual severe COVID-19 if she gets it as above average at 0.07%, but also a 99.85% chance she will not become infected at all.

### C. The urban Asian bus driver

Rajiv is a 35-year-old London bus driver of Indian origin. He is a fitness enthusiast and takes care to maintain a healthy weight. Despite that, he has type 1 diabetes. He gave up smoking last year. Rajiv does not observe any religion, and he lives in a comfortable home with no overcrowding. He takes care not to mix with other people too much outside of work, and he feels confident he will not become too ill even if he does catch COVID-19 because of his youth and his commitment to fitness. The model predicts his probability of eventually contracting severe COVID-19 at 0.04%, slightly higher than that of the general population. He is more likely than the average person to contract some degree of COVID-19, with a 99.73% chance of not contracting it at all.

### D. The African hospital orderly

Victor is a 29-year-old Oxford hospital orderly, Black, of Ghanaian origin. His weight is normal, he has never smoked and has no known medical conditions. He goes regularly to an evangelical church but otherwise avoids other people at the moment, although he has to use public transport to travel to work. He lives with a flatmate who works from home. He has been in contact with patients with COVID-19 symptoms at work although he does not work in a COVID-19 ward and has not knowingly been in close contact with a confirmed COVID-19 patient. The model predicts a 0.2% chance of severe COVID-19 – 10 times that of the general population - and only an 97.64% chance of not contracting the disease at all.

### E. The Chinese schoolgirl

Yun Chee is an 11-year-old Chinese schoolgirl living in London. She is of normal weight, has no underlying medical conditions, and is not religious. She had only been to school a few times in the past months until her school reopened in September. She has also started going on playdates and sleepovers with her friends. She lives in a small house with her parents and grandparents, with one bathroom and one kitchen. Yun Chee has a lower-than-average probability of 0.01% of contracting severe COVID-19, the model predicts, but almost double the average probability of eventually having it asymptomatically, at 0.13%. This is a risk that should be monitored closely and may need to be adjusted according to observational data as schools remain open.

## VII. CONCLUSION

A Bayesian network such as the one presented here could be a powerful tool for evaluating individual risk as countries around the world seek to lift lockdown measures without releasing a second wave of coronavirus. It is more nuanced than current health advice and more comprehensive than other privacy-preserving contact-tracing apps that have been proposed. Its efficacy does depend on a sense of social responsibility but it can also help breed that responsibility by heightening risk awareness.

Testing the network presents several challenges, and mainly consists of running numerous scenarios to validate the model against what data there is:

- There are as yet very few general-population studies of COVID-19, meaning that almost all the knowledge we have so far is based on hospital patients.
- The studies that do exist are observational, not experimental.
- Little is known about how many people have COVID-19 without symptoms, as has been noted in (Neil, et al., 2020).
- As soon as an observation is entered into the model, it no longer represents general population data.

However, the first three of these problems are relevant for any predictive model. In the circumstances, a causal probabilistic model that uses expert knowledge is likely to be a better tool than other types of statistical analysis.

There are a few studies emerging that, while still not general-population surveys, do include people who are not ill or infected and provide valuable material for cross-checking probabilities. One such example is (Poletti, et al., 2020), a study of 64,252 close contacts of 21,410 COVID-19 cases in Lombardy, Italy who were tested between February and April 2020. This study was aimed at discovering the probability of developing symptoms given infection, and was conditioned on age and sex. As testing becomes more prevalent and more such studies are possible, the Bayesian model can be further adjusted to reflect the latest state of knowledge.

## VIII. FUTURE WORK

The revised complete BN model, which includes both the extended background risks and infection possibility described in this report together with the extended set of symptoms described in (Butcher 2020), can be seen in the Appendix.

More work needs to be done on immunity possibility, test types and results, and prevalence and infectiousness of asymptomatic and pre-symptomatic COVID-19 as more evidence becomes available. In general, the model needs to be constantly updated to keep up with the fast-developing state of scientific knowledge.

The power of the Bayesian model could be more explicitly leveraged by including interventions such as “go to school”, “wear a mask” or “take public transport”.

If the network were to be used for policy-making rather than personal risk assessment, counterfactuals such as “close international borders”, “shut down pubs” or “postpone exams” could be added, taking advantage of another characteristic of Bayesian networks. Utility nodes could also be added - for example, to model the positive utility of recommencing education against the negative utility of increasing infection risks.

## Data Availability

Links to all data sources are provided in the document.

http://www.eecs.qmul.ac.uk/~norman/Models/

## APPENDIX

This appendix shows the full model, followed by the nodes on which I worked and details how I arrived at their probability tables.

### A. FULL MODEL

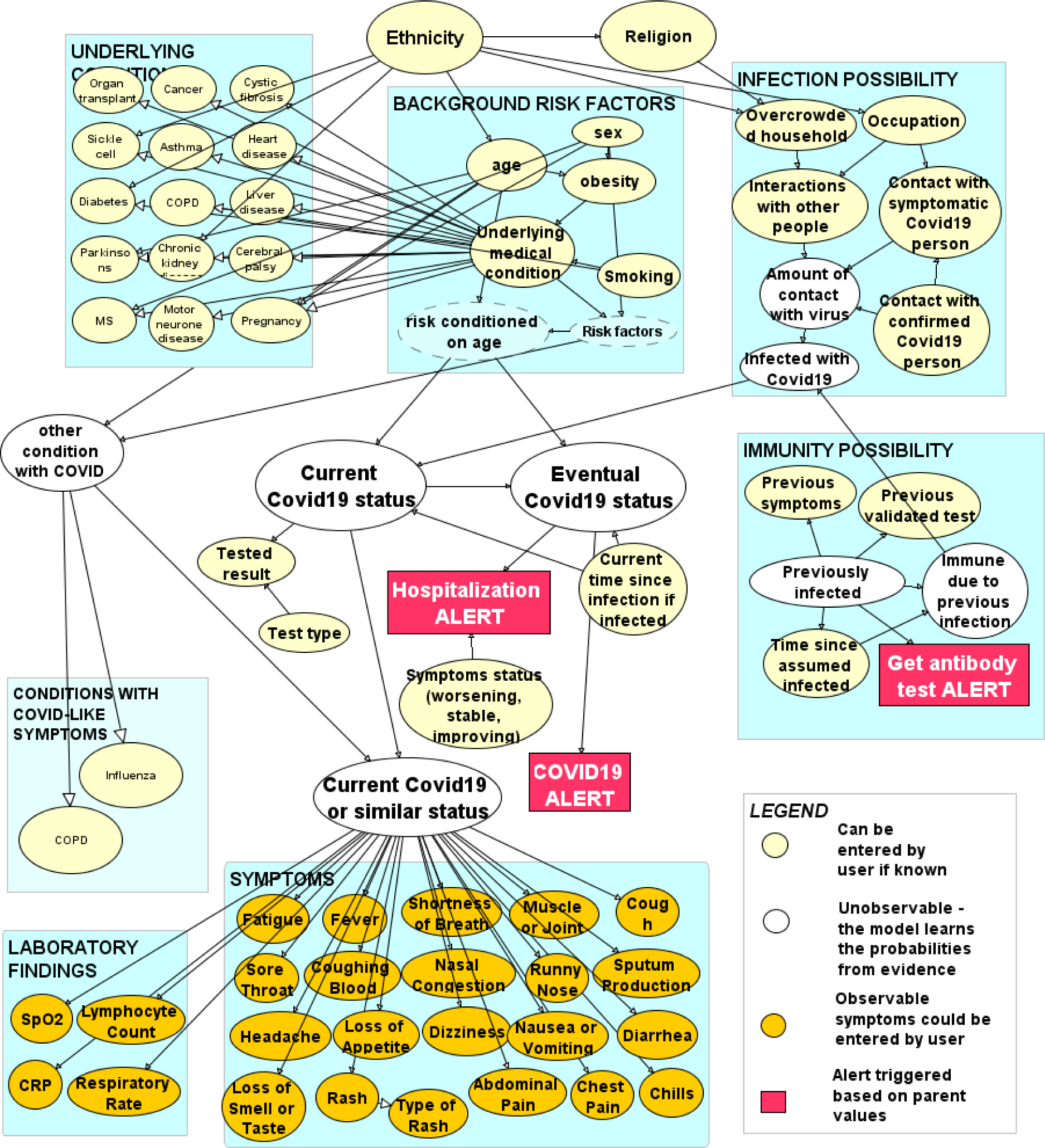

The model, entitled “CombinedModelForContactTracing.cmpx” can be downloaded at Norman Fenton’s home page http://www.eecs.qmul.ac.uk/~norman/Models/ and can be run using trial software from agenarisk.com.

### B. NODES

The nodes are presented in the following sections:

1. Ethnicity
2. Religion
3. Other major background risk factors
4. Underlying medical conditions
5. Risk factors (combined)
6. Risk conditioned on age
7. Infection risks
8. Outcomes

Other nodes were either inherited from an earlier version of the model or were worked on by my colleague Rachel Butcher.

#### 1. ETHNICITY

|  |  |
| --- | --- |
| White | 0.84815186 |
| Mixed/Multiple | 0.017982017 |
| Chinese | 0.006993007 |
| Other Asian | 0.07292707 |
| Black | 0.034965035 |
| Other | 0.018981019 |

These data were taken from the latest population estimates (2019) by the Office for National Statistics. https://www.ons.gov.uk/peoplepopulationandcommunity/populationandmigration/populationestimates/articles/researchreportonpopulationestimatesbyethnicgroupandreligion/2019-12-04.

They are only for England and Wales but I decided that extrapolating the proportions to the rest of the UK was preferable to taking the 2011 Census statistics for the whole of the UK.

I supplemented this with government statistics on the Chinese ethnic group. https://www.ethnicity-facts-figures.service.gov.uk/summaries/chinese-ethnic-group.

It seemed important to separate Chinese from other Asian groups because of the very different health and cultural profiles of Chinese and South Asian.

It would also have been useful to separate Black African and Black Caribbean, for the same reasons.

#### 2. RELIGION

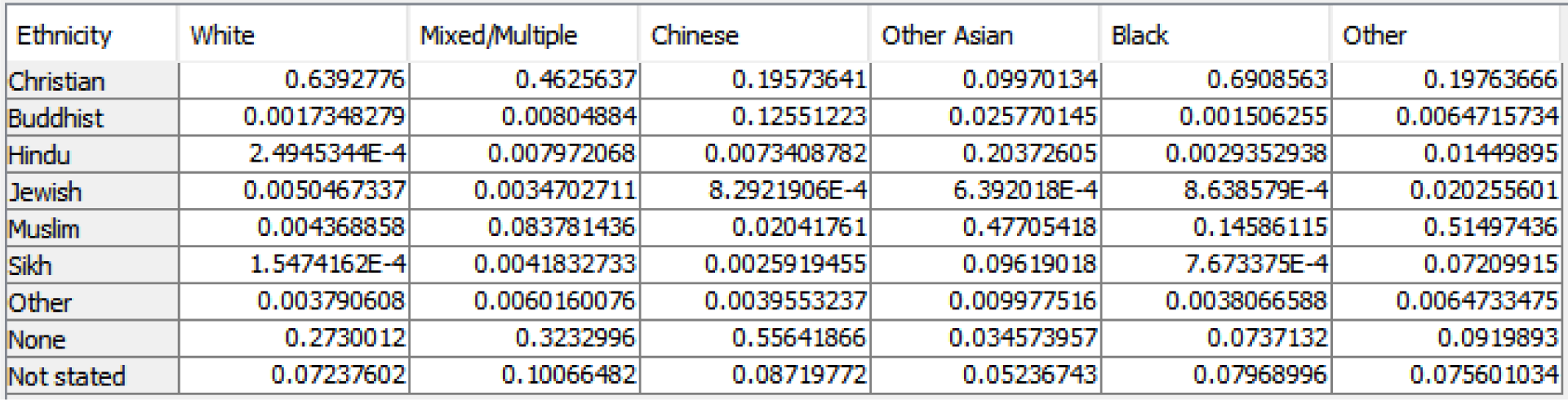

These data came from the ONS via a 2015 Freedom of Information request. https://www.ons.gov.uk/aboutus/transparencyandgovernance/freedomofinformationfoi/ethnicityandreligionbyage The absolute numbers in the xls file are normalised by the AgenaRisk software to add up to 1 in each column.

#### 3. BACKGROUND RISK FACTORS

##### 3.a. AGE

| Ethnicity | White | Mixed/Multiple | Chinese | Other Asian | Black | Other |
| --- | --- | --- | --- | --- | --- | --- |
| 75 and above | 0.08727237 | 0.012266416 | 0.020000458 | 0.021542113 | 0.025700176 | 0.017351551 |
| 65-74 | 0.095409095 | 0.016625285 | 0.029999414 | 0.035962854 | 0.036260583 | 0.028616488 |
| 55-64 | 0.12686937 | 0.028467821 | 0.07199962 | 0.06665908 | 0.04970856 | 0.0607597 |
| 35-54 | 0.2798486 | 0.16767396 | 0.24899973 | 0.2605109 | 0.3147982 | 0.2778235 |
| 25-34 | 0.124623135 | 0.14566073 | 0.21999995 | 0.21280235 | 0.16364022 | 0.22974086 |
| 16-24 | 0.11281764 | 0.1795067 | 0.29099992 | 0.14314783 | 0.13706492 | 0.14736845 |
| 0-15 | 0.1731598 | 0.4497991 | 0.11800092 | 0.2593749 | 0.27282736 | 0.23833945 |

I used data from the same FOI request as for the Religion node above.

The numbers needed some rearranging into the age groupings I had chosen. Here and in other similar situations I used this population-distribution calculator from the ONS to weight the results:

https://www.ons.gov.uk/peoplepopulationandcommunity/populationandmigration/populationestimates/articles/overviewoftheukpopulation/august2019

##### 3.b. SEX

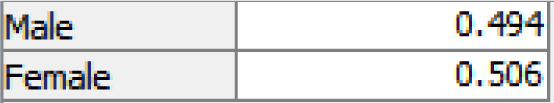

Data source: https://www.ons.gov.uk/peoplepopulationandcommunity/populationandmigration/populationestimates/bulletins/annualmidyearpopulationestimates/mid2019estimates

##### 3.c. OBESITY

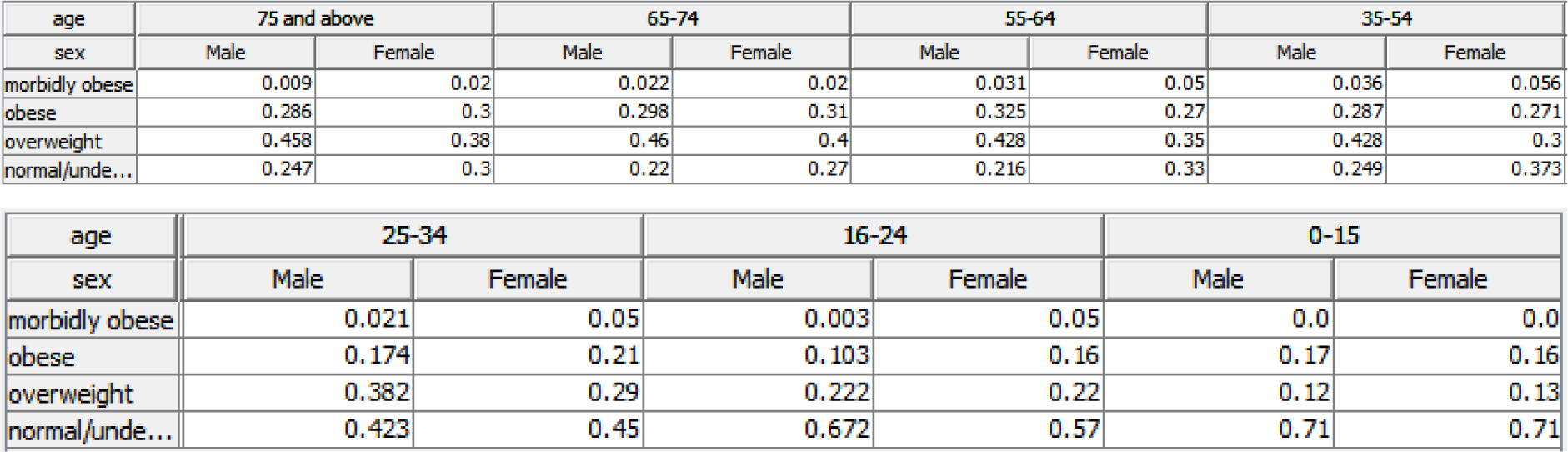

| age | 75 and above |  | 65-74 |  | 55-64 |  | 35-54 |  |
| --- | --- | --- | --- | --- | --- | --- | --- | --- |
| sex | Male | Female | Male | Female | Male | Female | Male | Female |
| morbidly obese | 0.009 | 0.02 | 0.022 | 0.02 | 0.031 | 0.05 | 0.036 | 0.056 |
| obese | 0.286 | 0.3 | 0.298 | 0.31 | 0.325 | 0.27 | 0.287 | 0.271 |
| overweight | 0.458 | 0.38 | 0.46 | 0.4 | 0.428 | 0.35 | 0.428 | 0.3 |
| normal/unde... | 0.247 | 0.3 | 0.22 | 0.27 | 0.216 | 0.33 | 0.249 | 0.373 |

| age | 25-34 |  | 16-24 |  | 0-15 |  |
| --- | --- | --- | --- | --- | --- | --- |
| sex | Male | Female | Male | Female | Male | Female |
| morbidly obese | 0.021 | 0.05 | 0.003 | 0.05 | 0.0 | 0.0 |
| obese | 0.174 | 0.21 | 0.103 | 0.16 | 0.17 | 0.16 |
| overweight | 0.382 | 0.29 | 0.222 | 0.22 | 0.12 | 0.13 |
| normal/unde... | 0.423 | 0.45 | 0.672 | 0.57 | 0.71 | 0.71 |

These data were taken from the NHS Health Survey for England 2017. (I could not find such detailed data in more recent surveys.)

https://digital.nhs.uk/data-and-information/publications/statistical/health-survey-for-england/2017 There were separate government figures available on obesity by ethnicity:

https://www.ethnicity-facts-figures.service.gov.uk/health/diet-and-exercise/overweight-adults/latest

But I was unable to find data on obesity by all three factors. I decided in favour of age and sex because a quick Excel analysis suggested they were bigger influencers of obesity than ethnicity. I was also able to capture some of the relationship between ethnicity and obesity by conditioning diabetes on ethnicity (see Diabetes section below).

##### 3.d. SMOKING

|  |  |
| --- | --- |
| current | 0.134 |
| previous | 0.352 |
| never | 0.514 |

Data are from here: https://www.ons.gov.uk/peoplepopulationandcommunity/healthandsocialcare/healthandlifeexpectancies/bulletins/adultsmokinghabitsingreatbritain/2019#data-on-smokers-who-have-quit-and-smokers-who-intend-to-quit-great-britain-1974-to-2019

The numbers are for adults aged 16 and above. I decided to assume there were no smokers aged below 16, given that numbers would be small and risks at that age, in terms of indicating an underlying health condition, would be minimal.

##### 3.e. UNDERLYING MEDICAL CONDITION

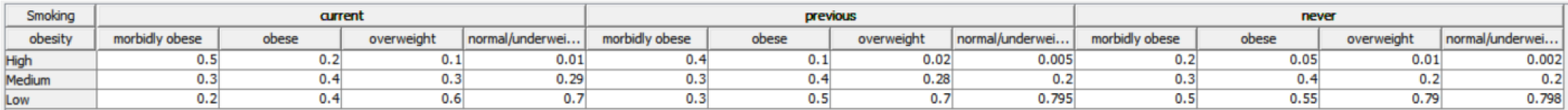

These are rough approximations conditioned on smoking and obesity status. The probabilities are altered by backward inference when an observation is made for any specific medical condition.

#### 4. UNDERLYING CONDITIONS

I took the risk classification from here: https://www.nhs.uk/conditions/coronavirus-covid-19/people-at-higher-risk/whos-at-higher-risk-from-coronavirus/

For each underlying condition, I first took the classification of high- or medium-risk, summed the total high-risk and medium-risk patients, and then calculated how probable this particular condition was given an overall rating of high or medium for “underlying medical condition”.

|  |  |  |  |  |  |  |  |  |  |  |  |  |
| --- | --- | --- | --- | --- | --- | --- | --- | --- | --- | --- | --- | --- |
| HIGH RISK | organ | cancer | cystic | sickle | TOTAL HIGH |  |  |  |  |  |  |  |
| number of cases | 53000 | 2900000 | 10500 | 15000 | 2978500 |  |  |  |  |  |  |  |
| % of population | 0% | 4% | 0% | - |  |  |  |  |  |  |  |  |
| % of high risk | 1.80% | 97.30% | 0.40% | 0.50% |  |  |  |  |  |  |  |  |
| % of all underlying |  |  |  |  |  |  |  |  |  |  |  |  |
| MODERATE RISK | asthma | COPD | heart | diabetes | kidney | liver | Parkinson's | motor | MS | cerebral | pregnant | TOTAL MODERATE |
| number of cases | 5400000 | 1500000 | 2300000 | 4700000 | 3000000 | 9300 | 145000 | 5000 | 130000 | 30000 | 549000 | 17768300 |
| % of population | 8% | 2% | 4% | 7% | 5% | 0% | 0% | 0% | 0% | 0% | 1% |  |
| % of moderate risk | 30.39% | 8.44% | 12.94% | 26.45% | 16.88% | 0.05% | 0.82% | 0.03% | 0.73% | 0.17% | 3.09% |  |
| % of all underlying | 26% | 7.23% | 11.10% | 22.70% | 14.50% | 0.04% | 0.70% | 0.02% | 0.63% | 0.14% | 2.65% |  |

##### 4.a. ORGAN TRANSPLANT

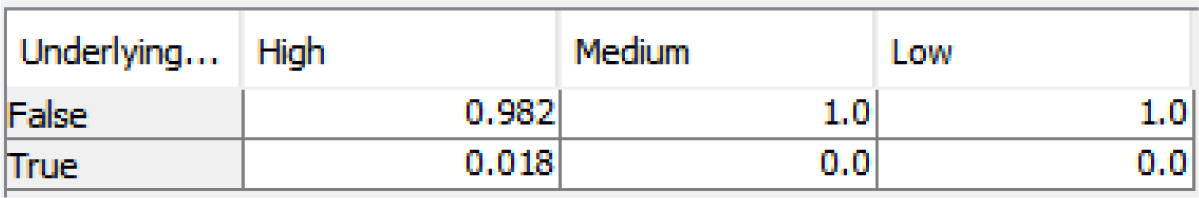

| Underlying... | High | Medium | Low |
| --- | --- | --- | --- |
| False | 0.982 | 1.0 | 1.0 |
| True | 0.018 | 0.0 | 0.0 |

https://www.organdonation.nhs.uk/get-involved/news/more-than-50-000-now-alive-thanks-to-organ-donations/

##### 4.b. CANCER

| Underlying... | High | Medium | Low |
| --- | --- | --- | --- |
| False | 0.027 | 1.0 | 1.0 |
| True | 0.973 | 0.0 | 0.0 |

https://www.macmillan.org.uk/_images/cancer-statistics-factsheet_tcm9-260514.pdf

##### 4.c. CYSTIC FIBROSIS

| Underlying... | High | Medium | Low |
| --- | --- | --- | --- |
| False | 0.996 | 1.0 | 1.0 |
| True | 0.004 | 0.0 | 0.0 |

https://www.cysticfibrosis.org.uk/what-is-cystic-fibrosis/faqs#:~:text=Around%2010%2C500%20people%20in%20the,100%2C000%20people%20in%20the%20world.

##### 4.d. SICKLE CELL DISEASE

| Underlying ... | High |  |  |  |  |  |
| --- | --- | --- | --- | --- | --- | --- |
| Ethnicity | White | Mixed/Multiple | Chinese | Other Asian | Black | Other |
| False | 0.99822 | 0.99822 | 0.99822 | 0.99822 | 0.91 | 0.99822 |
| True | 0.00178 | 0.00178 | 0.00178 | 0.00178 | 0.09 | 0.00178 |

| Underlying ... | Medium |  |  |  |  |  |
| --- | --- | --- | --- | --- | --- | --- |
| Ethnicity | White | Mixed/Multiple | Chinese | Other Asian | Black | Other |
| False | 1.0 | 1.0 | 1.0 | 1.0 | 1.0 | 1.0 |
| True | 0.0 | 0.0 | 0.0 | 0.0 | 0.0 | 0.0 |

| Underlying ... | Low |  |  |  |  |  |
| --- | --- | --- | --- | --- | --- | --- |
| Ethnicity | White | Mixed/Multiple | Chinese | Other Asian | Black | Other |
| False | 1.0 | 1.0 | 1.0 | 1.0 | 1.0 | 1.0 |
| True | 0.0 | 0.0 | 0.0 | 0.0 | 0.0 | 0.0 |

https://www.sicklecellsociety.org/about-sickle-cell/#:~:text=4%20Approximately%2015%2C000%20people%20in,in%20the%20UK%20every%20year.&text=7%20Children%20with%20SCD%20are,ages%20of%202%20and%2016

##### 4.e. ASTHMA

| Underlying... | High | Medium | Low |
| --- | --- | --- | --- |
| False | 0.74 | 0.74 | 1.0 |
| True | 0.26 | 0.26 | 0.0 |

https://www.asthma.org.uk/about/media/facts-and-statistics/

##### 4.f. HEART DISEASE

| Underlying... | High | Medium | Low |
| --- | --- | --- | --- |
| False | 0.889 | 0.889 | 1.0 |
| True | 0.111 | 0.111 | 0.0 |

https://www.bhf.org.uk/what-we-do/our-research/heart-statistics

##### 4.g. DIABETES

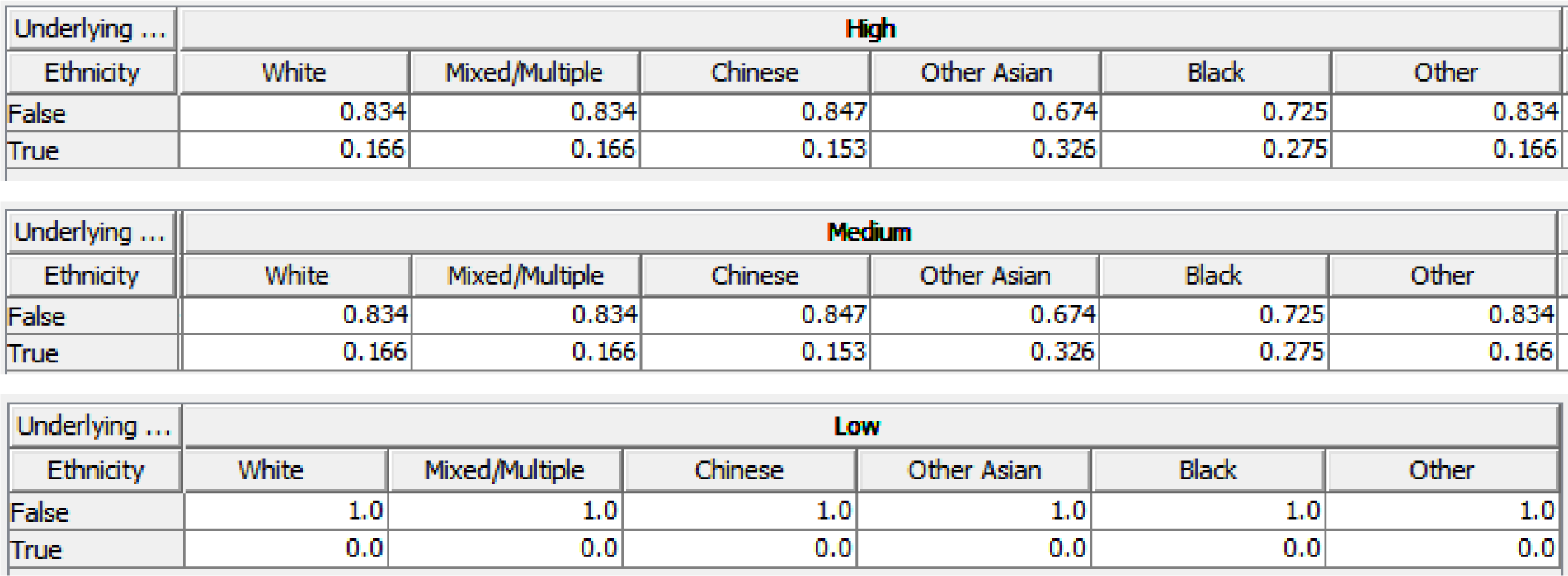

https://www.diabetes.org.uk/resources-s3/2017-11/diabetes_in_the_uk_2010.pdf

##### 4.h. COPD

| Underlying ... | High |  |  | Medium |  |  | Low |  |  |
| --- | --- | --- | --- | --- | --- | --- | --- | --- | --- |
| Smoking | current | previous | never | current | previous | never | current | previous | never |
| False | 0.935 | 0.935 | 0.993 | 0.935 | 0.935 | 0.993 | 1.0 | 1.0 | 1.0 |
| True | 0.065 | 0.065 | 0.0069999998 | 0.065 | 0.065 | 0.0069999998 | 0.0 | 0.0 | 0.0 |

https://www.hse.gov.uk/statistics/causdis/copd.pdf https://www.nhs.uk/conditions/chronic-obstructive-pulmonary-disease-copd/causes/

##### 4.i. LIVER DISEASE

| Underlying... | High | Medium | Low |
| --- | --- | --- | --- |
| False | 0.9996 | 0.9996 | 1.0 |
| True | 4.0E-4 | 4.0E-4 | 0.0 |

https://www.statista.com/statistics/1036415/prevalence-of-cirrhosis-in-the-uk-by-gender/

##### 4.j. PARKINSON’S DISEASE

4.j. PARKINSON'S DISEASE
| Underlying ... | High |  |  |  |  |  |  |
| --- | --- | --- | --- | --- | --- | --- | --- |
| age | 75 and above | 65-74 | 55-64 | 35-54 | 25-34 | 16-24 | 0-15 |
| False | 0.974 | 0.9931 | 0.9994 | 0.99998 | 1.0 | 1.0 | 1.0 |
| True | 0.025999999 | 0.0069 | 5.9999997E-4 | 2.0000001E-5 | 0.0 | 0.0 | 0.0 |

| Medium |  |  |  |  |  |  |
| --- | --- | --- | --- | --- | --- | --- |
| 75 and above | 65-74 | 55-64 | 35-54 | 25-34 | 16-24 | 0-15 |
| 0.974 | 0.9931 | 0.9994 | 0.99998 | 1.0 | 1.0 | 1.0 |
| 0.025999999 | 0.0069 | 5.9999997E-4 | 2.0000001E-5 | 0.0 | 0.0 | 0.0 |

| Low |  |  |  |  |  |  |
| --- | --- | --- | --- | --- | --- | --- |
| 75 and above | 65-74 | 55-64 | 35-54 | 25-34 | 16-24 | 0-15 |
| 1.0 | 1.0 | 1.0 | 1.0 | 1.0 | 1.0 | 1.0 |
| 0.0 | 0.0 | 0.0 | 0.0 | 0.0 | 0.0 | 0.0 |

https://www.parkinsons.org.uk/professionals/news/parkinsons-diagnoses-rise-uk#:~:text=The%20number%20of%20people%20diagnosed,350%20adults%20in%20the%20UK

##### 4.k. CHRONIC KIDNEY DISEASE

| Underlying ... | High |  |  |  |  |  |
| --- | --- | --- | --- | --- | --- | --- |
| Ethnicity | White | Mixed/Multiple | Chinese | Other Asian | Black | Other |
| False | 0.862 | 0.938 | 0.876 | 0.8 | 0.736 | 0.938 |
| True | 0.138 | 0.062 | 0.124 | 0.2 | 0.264 | 0.062 |

| Medium |  |  |  |  |  |
| --- | --- | --- | --- | --- | --- |
| White | Mixed/Multiple | Chinese | Other Asian | Black | Other |
| 0.862 | 0.938 | 0.876 | 0.8 | 0.736 | 0.938 |
| 0.138 | 0.062 | 0.124 | 0.2 | 0.264 | 0.062 |

| Low |  |  |  |  |  |
| --- | --- | --- | --- | --- | --- |
| White | Mixed/Multiple | Chinese | Other Asian | Black | Other |
| 1.0 | 1.0 | 1.0 | 1.0 | 1.0 | 1.0 |
| 0.0 | 0.0 | 0.0 | 0.0 | 0.0 | 0.0 |

https://www.renalreg.org/wp-content/uploads/2014/09/06-Chap-06.pdf

##### 4.l. CEREBRAL PALSY

| Underlying... | High | Medium | Low |
| --- | --- | --- | --- |
| False | 0.9986 | 0.9986 | 1.0 |
| True | 0.0014 | 0.0014 | 0.0 |

https://thepacecentre.org/information-centre/stats-facts/

##### 4.m. MULTIPLE SCLEROSIS

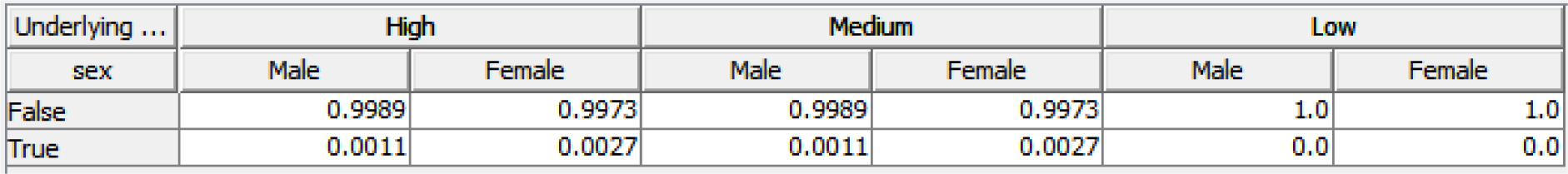

https://www.gov.uk/government/publications/multiple-sclerosis-prevalence-incidence-and-smoking-status/multiple-sclerosis-prevalence-incidence-and-smoking-status-data-briefing

##### 4.n. MOTOR NEURONE DISEASE

| Underlying... | High | Medium | Low |
| --- | --- | --- | --- |
| False | 0.9998 | 0.9998 | 1.0 |
| True | 2.0E-4 | 2.0E-4 | 0.0 |

https://www.nhsinform.scot/illnesses-and-conditions/brain-nerves-and-spinal-cord/motor-neurone-disease-mnd#:~:text=Motor%20neurone%20disease%20is%20a%20rare%20condition%20that%20affects%20around,although%20this%20is%20extremely%20rare.

##### 4.o. PREGNANCY

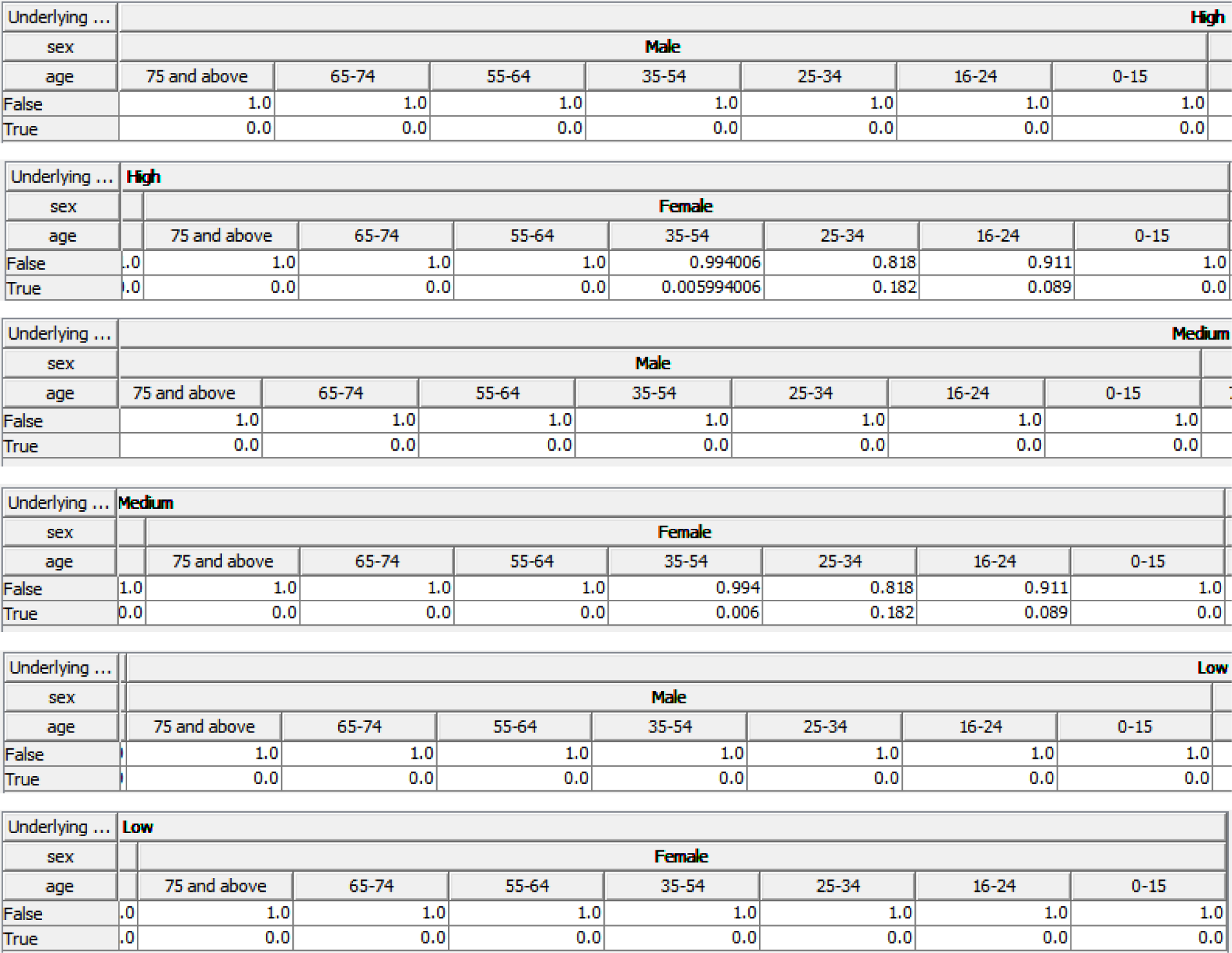

https://www.statista.com/statistics/297718/conception-rate-per-thousand-women-in-england-and-wales-by-age/

#### 5 RISK FACTORS

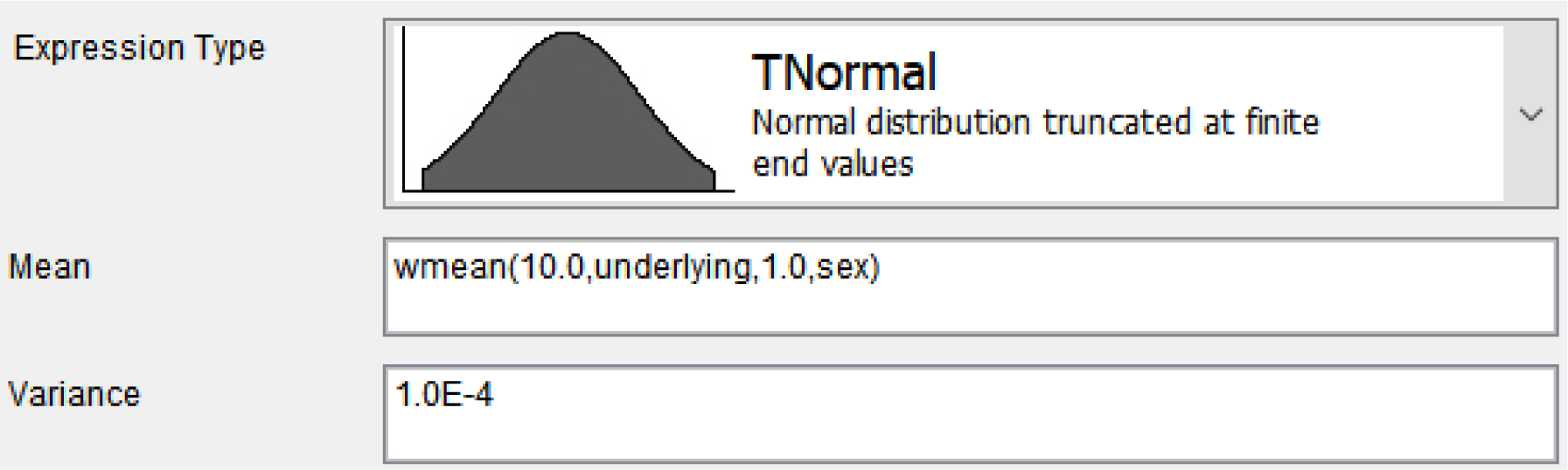

5. RISK FACTORS
|  |  |
| --- | --- |
| Expression Type | **TNormal**<br>Normal distribution truncated at finite end values |
| Mean | wmean(10.0,underlying,1.0,sex) |
| Variance | 1.0E-4 |

#### 6 RISK CONDITIONED ON AGE

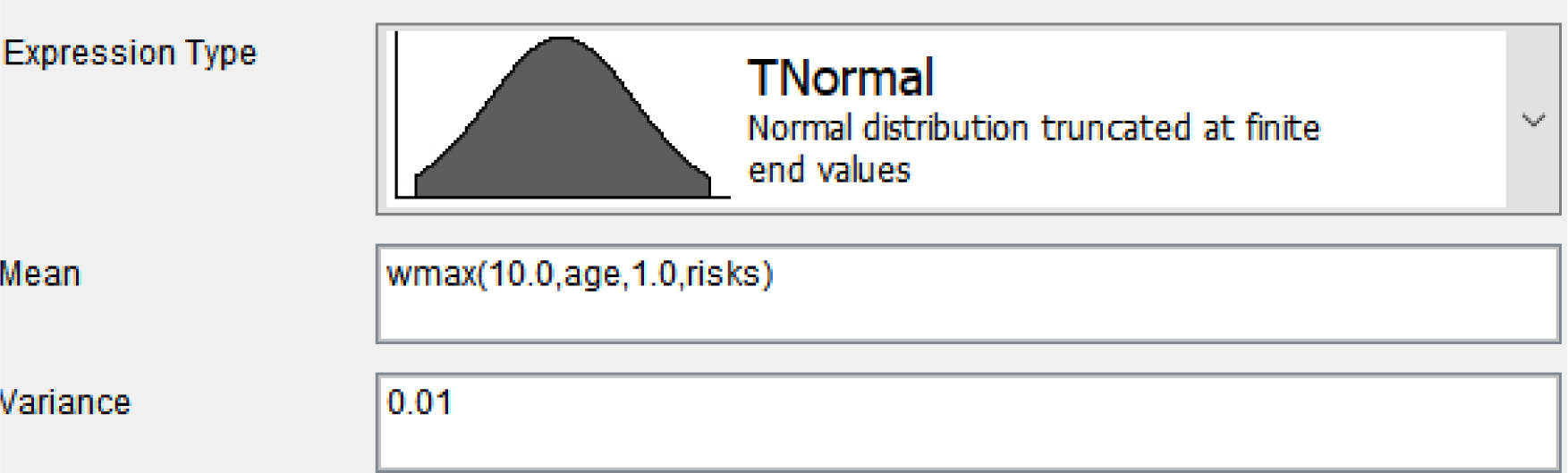

#### 7 INFECTION RISKS

##### 7.a. OCCUPATION

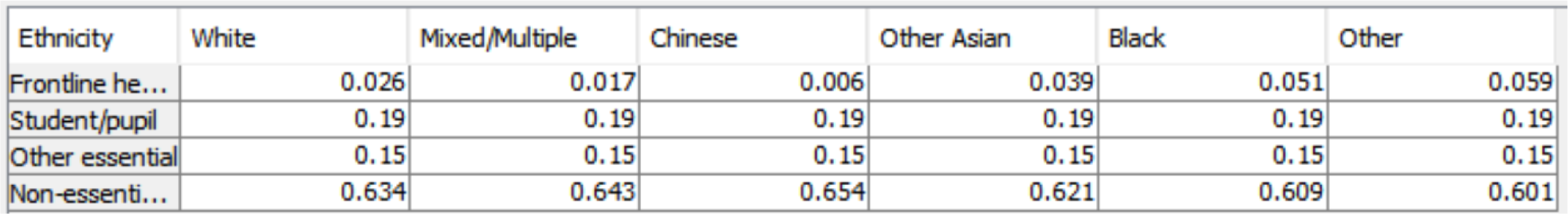

7. INFECTION RISKS 7.a. OCCUPATION
| Ethnicity | White | Mixed/Multiple | Chinese | Other Asian | Black | Other |
| --- | --- | --- | --- | --- | --- | --- |
| Frontline he... | 0.026 | 0.017 | 0.006 | 0.039 | 0.051 | 0.059 |
| Student/pupil | 0.19 | 0.19 | 0.19 | 0.19 | 0.19 | 0.19 |
| Other essential | 0.15 | 0.15 | 0.15 | 0.15 | 0.15 | 0.15 |
| Non-essenti... | 0.634 | 0.643 | 0.654 | 0.621 | 0.609 | 0.601 |

https://www.ethnicity-facts-figures.service.gov.uk/workforce-and-business/workforce-diversity/nhs-workforce/latest https://assets.publishing.service.gov.uk/government/uploads/system/uploads/attachment_data/file/908434/Disparities_in_the_risk_and_outcomes_of_COVID_August_2020_update.pdf

https://assets.publishing.service.gov.uk/government/uploads/system/uploads/attachment_data/file/812539/Schools_Pupils_and_their_Characteristics_2019_Main_Text.pdf

https://commonslibrary.parliament.uk/research-briefings/cbp-7857/#:~:text=In%20academic%20year%202018%2F19,at%20UK%20higher%20education%20institutions.&text=There%20were%20just%20over%20700%2C000,2019%20and%20541%2C000%20were%20accepted.

##### 7.b. OVERCROWDED HOUSEHOLD

| Ethnicity | White |  |  |  |  |  |  |  |  |
| --- | --- | --- | --- | --- | --- | --- | --- | --- | --- |
| Religion | Christian | Buddhist | Hindu | Jewish | Muslim | Sikh | Other | None | Not stated |
| Yes | 0.02 | 0.02 | 0.02 | 0.09 | 0.02 | 0.02 | 0.02 | 0.02 | 0.02 |
| No | 0.98 | 0.98 | 0.98 | 0.91 | 0.98 | 0.98 | 0.98 | 0.98 | 0.98 |

| Ethnicity | Mixed/Multiple |  |  |  |  |  |  |  |  |
| --- | --- | --- | --- | --- | --- | --- | --- | --- | --- |
| Religion | Christian | Buddhist | Hindu | Jewish | Muslim | Sikh | Other | None | Not stated |
| Yes | 0.058000002 | 0.058000002 | 0.058000002 | 0.09 | 0.058000002 | 0.058000002 | 0.058000002 | 0.058000002 | 0.058000002 |
| No | 0.942 | 0.942 | 0.942 | 0.91 | 0.942 | 0.942 | 0.942 | 0.942 | 0.942 |

| Ethnicity | Chinese |  |  |  |  |  |  |  |  |
| --- | --- | --- | --- | --- | --- | --- | --- | --- | --- |
| Religion | Christian | Buddhist | Hindu | Jewish | Muslim | Sikh | Other | None | Not stated |
| Yes | 0.07 | 0.07 | 0.07 | 0.09 | 0.07 | 0.07 | 0.07 | 0.07 | 0.07 |
| No | 0.93 | 0.93 | 0.93 | 0.91 | 0.93 | 0.93 | 0.93 | 0.93 | 0.93 |

| Ethnicity | Other Asian |  |  |  |  |  |  |  |  |
| --- | --- | --- | --- | --- | --- | --- | --- | --- | --- |
| Religion | Christian | Buddhist | Hindu | Jewish | Muslim | Sikh | Other | None | Not stated |
| Yes | 0.124 | 0.124 | 0.124 | 0.09 | 0.124 | 0.124 | 0.124 | 0.124 | 0.124 |
| No | 0.876 | 0.876 | 0.876 | 0.91 | 0.876 | 0.876 | 0.876 | 0.876 | 0.876 |

| Ethnicity | Black |  |  |  |  |  |  |  |  |
| --- | --- | --- | --- | --- | --- | --- | --- | --- | --- |
| Religion | Christian | Buddhist | Hindu | Jewish | Muslim | Sikh | Other | None | Not stated |
| Yes | 0.121999994 | 0.121999994 | 0.121999994 | 0.09 | 0.121999994 | 0.121999994 | 0.121999994 | 0.121999994 | 0.121999994 |
| No | 0.878 | 0.878 | 0.878 | 0.91 | 0.878 | 0.878 | 0.878 | 0.878 | 0.878 |

| Ethnicity | Other |  |  |  |  |  |  |  |  |
| --- | --- | --- | --- | --- | --- | --- | --- | --- | --- |
| Religion | Christian | Buddhist | Hindu | Jewish | Muslim | Sikh | Other | None | Not stated |
| Yes | 0.084 | 0.084 | 0.084 | 0.09 | 0.084 | 0.084 | 0.084 | 0.084 | 0.084 |
| No | 0.916 | 0.916 | 0.916 | 0.91 | 0.916 | 0.916 | 0.916 | 0.916 | 0.916 |

https://www.ethnicity-facts-figures.service.gov.uk/housing/housing-conditions/overcrowded-households/latest https://www.jpr.org.uk/documents/JPR_Census_Jewish_families_and_Jewish_households_report_March_2015.pdf

##### 7.c. CONTACT WITH SYMPTOMATIC COVID-19 PERSON

| Contact with confirmed ... | Yes |  |  |  | No |  |  |  |
| --- | --- | --- | --- | --- | --- | --- | --- | --- |
| Occupation | Frontline healthc... | Student/pupil | Other essential | Non-essential/no... | Frontline healthc... | Student/pupil | Other essential | Non-essential/no... |
| Yes | 1.0 | 0.9 | 1.0 | 1.0 | 0.1 | 0.02 | 0.015 | 0.01 |
| No | 0.0 | 0.1 | 0.0 | 0.0 | 0.9 | 0.98 | 0.985 | 0.99 |

##### 7.d. CONTACT WITH CONFIRMED COVID-19 PERSON

|  |  |
| --- | --- |
| Yes | 0.02 |
| No | 0.98 |

https://www.gov.uk/government/publications/nhs-test-and-trace-england-and-coronavirus-testing-uk-statistics-6-august-to-12-august-2020/weekly-statistics-for-nhs-test-and-trace-england-and-coronavirus-testing-uk-6-august-to-12-august

##### 7.e. INTERACTIONS WITH OTHER PEOPLE

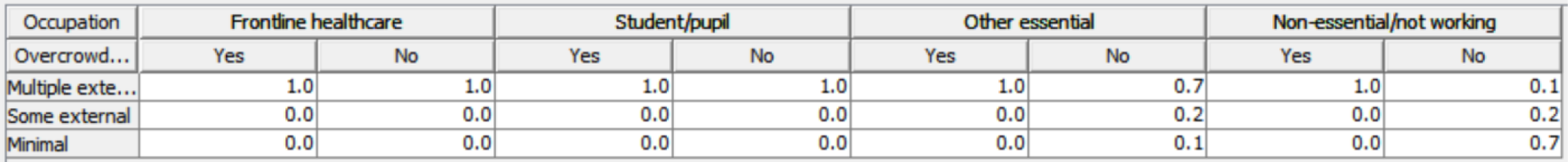

##### 7.f. AMOUNT OF CONTACT WITH VIRUS

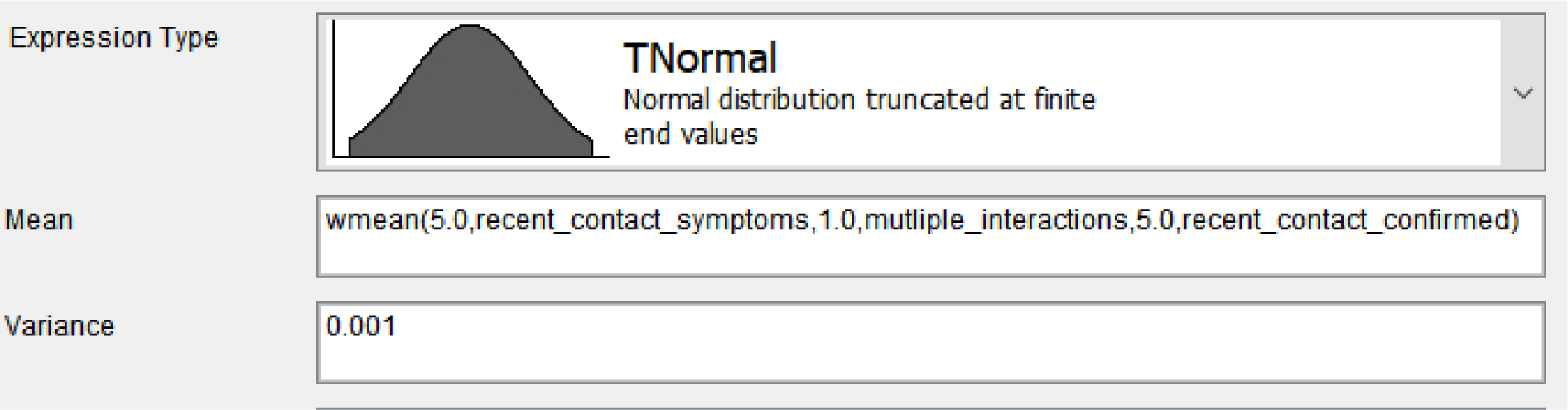

#### 8 OUTCOMES

##### 8.a. INFECTED WITH COVID-19

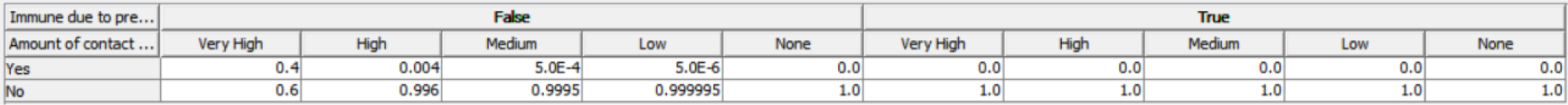

Government community infection survey pilot: https://www.ons.gov.uk/peoplepopulationandcommunity/healthandsocialcare/conditionsanddiseases/bulletins/coronaviruscovid19infectionsurveypilot/englandandwales21august2020

##### 8.b. CURRENT TIME SINCE INFECTION IF INFECTED

|  |  |
| --- | --- |
| > 5 days | 0.5 |
| <= 5 days | 0.5 |

This on how long COVID-19 remains infectious: https://covid.joinzoe.com/post/covid-long-term And this on incubation period: https://www.acpjournals.org/doi/10.7326/M20-0504

##### 8.c. CURRENT COVID-19 STATUS

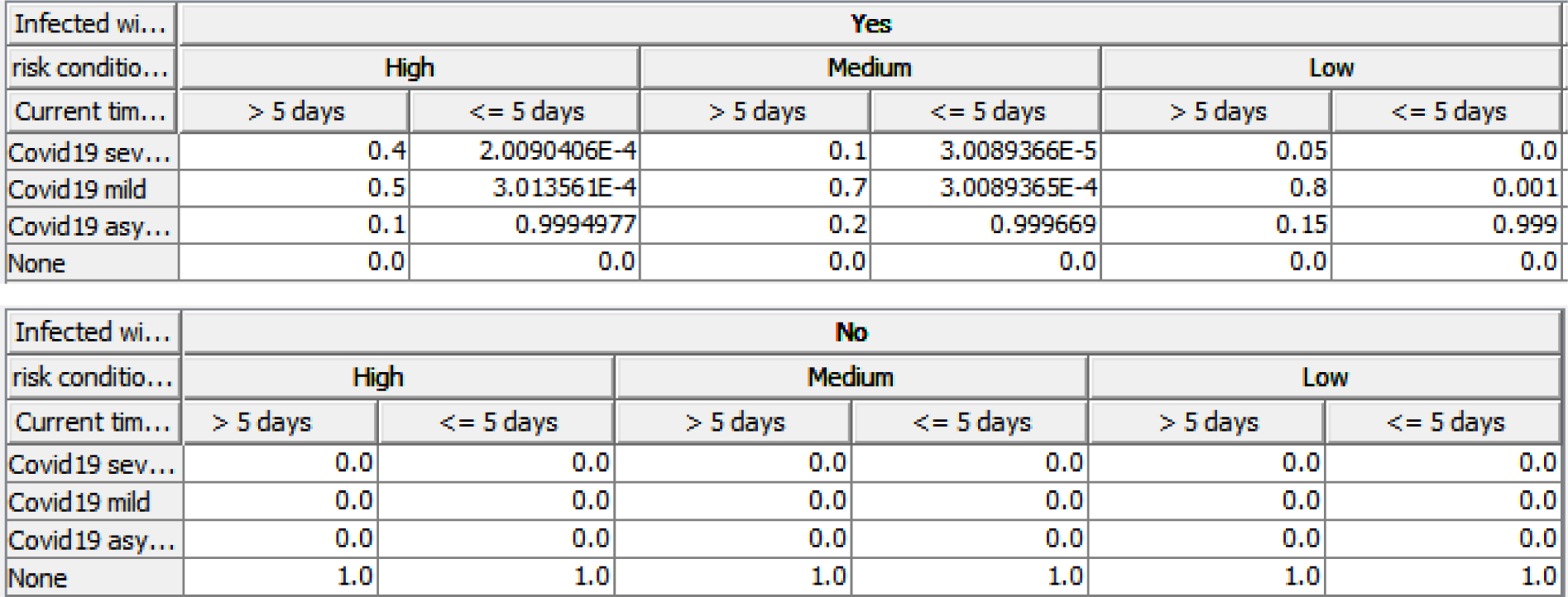

https://www.ons.gov.uk/peoplepopulationandcommunity/healthandsocialcare/conditionsanddiseases/bulletins/coronaviruscovid19infectionsurveypilot/englandandwales21august2020

##### 8.d. EVENTUAL COVID-19 STATUS

| Current Covid19 status | Covid19 severe |  |  |  |  |  |
| --- | --- | --- | --- | --- | --- | --- |
| risk conditioned on age | High |  | Medium |  | Low |  |
| Current time since inf... | > 5 days | <= 5 days | > 5 days | <= 5 days | > 5 days | <= 5 days |
| Covid19 severe | 1.0 | 1.0 | 1.0 | 1.0 | 1.0 | 1.0 |
| Covid19 mild | 0.0 | 0.0 | 0.0 | 0.0 | 0.0 | 0.0 |
| Covid19 asymptomatic | 0.0 | 0.0 | 0.0 | 0.0 | 0.0 | 0.0 |
| None | 0.0 | 0.0 | 0.0 | 0.0 | 0.0 | 0.0 |

| Current Covid19 status | Covid19 mild |  |  |  |  |  |
| --- | --- | --- | --- | --- | --- | --- |
| risk conditioned on age | High |  | Medium |  | Low |  |
| Current time since inf... | > 5 days | <= 5 days | > 5 days | <= 5 days | > 5 days | <= 5 days |
| Covid19 severe | 0.2 | 0.9 | 0.1 | 0.5 | 0.01 | 0.1 |
| Covid19 mild | 0.8 | 0.1 | 0.9 | 0.5 | 0.99 | 0.9 |
| Covid19 asymptomatic | 0.0 | 0.0 | 0.0 | 0.0 | 0.0 | 0.0 |
| None | 0.0 | 0.0 | 0.0 | 0.0 | 0.0 | 0.0 |

| Current Covid19 status | Covid19 asymptomatic |  |  |  |  |  |
| --- | --- | --- | --- | --- | --- | --- |
| risk conditioned on age | High |  | Medium |  | Low |  |
| Current time since inf... | > 5 days | <= 5 days | > 5 days | <= 5 days | > 5 days | <= 5 days |
| Covid19 severe | 0.1 | 0.7 | 0.01 | 0.1 | 0.0 | 0.05 |
| Covid19 mild | 0.5 | 0.25 | 0.29 | 0.7 | 0.2 | 0.1 |
| Covid19 asymptomatic | 0.4 | 0.05 | 0.7 | 0.2 | 0.8 | 0.85 |
| None | 0.0 | 0.0 | 0.0 | 0.0 | 0.0 | 0.0 |

| Current Covid19 status | None |  |  |  |  |  |
| --- | --- | --- | --- | --- | --- | --- |
| risk conditioned on age | High |  | Medium |  | Low |  |
| Current time since inf... | > 5 days | <= 5 days | > 5 days | <= 5 days | > 5 days | <= 5 days |
| Covid19 severe | 0.0 | 0.0 | 0.0 | 0.0 | 0.0 | 0.0 |
| Covid19 mild | 0.0 | 0.0 | 0.0 | 0.0 | 0.0 | 0.0 |
| Covid19 asymptomatic | 0.0 | 0.0 | 0.0 | 0.0 | 0.0 | 0.0 |
| None | 1.0 | 1.0 | 1.0 | 1.0 | 1.0 | 1.0 |

## Notes

### Competing Interest Statement

The authors have declared no competing interest.

### Funding Statement

No external funding was received

### Author Declarations

Queen Mary, University of London

